# Does Data Preprocessing Affect Tree-Based Super Learners? An Investigation of Ensemble Optimization and Oracle Properties in Clinical Classification

**DOI:** 10.64898/2026.08.20.26360880

**Authors:** Richmond Darko, Dorothy Dwumah, Kenneth Agyapong Saka, Yaw Agyenim-Boateng, Ronald Darko Anim, Jolly Wisdom Jakper, Rockson Owusu-Ansah, Nana Kwabena Owusu-Ansah

## Abstract

Machine learning workflows frequently incorporate data preprocessing to enhance predictive performance. However, the need for Super Learner ensembles made up only of preprocessing-invariant tree-based algorithms remains unexplored. Using three benchmark clinical classification datasets, this study examined how preprocessing affected the Super Learner’s prediction performance, learner weight distribution, and oracle behavior. The Heart Disease (207 observations), Indian Liver Patient Dataset (583 observations), and Pima Indians Diabetes (768 observations) datasets were used to create a Super Learner ensemble model that included Classification and Regression Trees (CART), Random Forest, Ranger, and Extreme Gradient Boosting (XGBoost). Models were evaluated under raw and preprocessed data conditions using repeated cross-validation. Predictive performance was assessed using the area under the receiver operating characteristic curve (AUC), Matthews correlation coefficient (MCC), and Brier score. Learner weight allocation and Oracle Gap were compared using paired Wilcoxon signed-rank tests with Benjamini–Hochberg adjustment. Preprocessing produced negligible changes in predictive performance for the Heart Disease and Pima datasets. For the ILPD dataset, preprocessing significantly improved AUC (0.746 to 0.752; adjusted *p* = 0.0017) and reduced the Brier score (0.177 to 0.175; adjusted *p <* 0.001). Learner weights remained largely stable, although Random Forest replaced Ranger as the dominant learner for the Heart Disease dataset. Oracle Gaps remained extremely small (*<*0.002) across all datasets and did not differ significantly between preprocessing conditions. Preprocessing provides limited benefit for Super Learner ensembles composed of preprocessing-invariant learners and does not materially alter their oracle behavior. Preprocessing decisions should therefore be guided by dataset characteristics rather than adopted as a universal modelling practice.

## 1. Introduction

Machine learning techniques are being used more often in healthcare for diagnosis and prognosis due to the increasing availability of biological and clinical data. Accurate prediction models that can identify high-risk patients and optimize treatment protocols are in great demand as a result of this surge [1]. Because of their interpretability, traditional statistical techniques like logistic regression have been fundamental to clinical prediction modeling; however, they are unable to handle the complexity that exists in modern healthcare data, which frequently has high dimensionality and nonlinear relationships[2].

Machine learning algorithms, such as decision trees, random forests, and neural networks, demonstrated efficacy in managing this complexity and enhancing prediction accuracy in a variety of medical disorders, such as cancer, diabetes, and cardiovascular diseases[3]. However, no single algorithm consistently outperforms the others, as their performance depends on data characteristics like sample size and distribution. This has led to the development of ensemble learning methods that combine multiple algorithms to enhance predictive performance and address the issues in selecting the most appropriate approach for different datasets[4, 5].

### 1.1. Evolution from Single Learners to Ensemble Learning

Numerous algorithms, each with unique advantages and disadvantages, have been developed as a result of the growth of machine learning in predictive modeling. While some are better at handling complicated interactions and nonlinear patterns, others are better at linear relationships. This inconsistency in performance across different datasets underscores the “No Free Lunch” theorem, which states that no single algorithm can outperform all others in every scenario[6–8].

The performance of algorithms in the healthcare industry is extremely dependent on the context due to the difficulties of different sample sizes, feature dimensionality, class imbalances, and noise. Relying on a single model may result in less generalizability and unstable results[5]. Researchers have used ensemble learning techniques, which aggregate predictions from several models to improve accuracy and stability, to solve these problems. Predictive performance has been greatly enhanced by strategies like bagging and boosting. While Random Forests further improve this by using random feature selection, Bagging generates several bootstrap samples and aggregates predictions. In order to achieve high accuracy, boosting techniques like AdaBoost and gradient boosting build models progressively, concentrating on data points that prior models incorrectly classified[9].

By combining predictions from several models, ensemble learning has become a successful method for getting beyond the shortcomings of individual learning algorithms. According to theoretical considerations, variance reduction and stronger reliability across datasets are the main ways that ensemble techniques improve predictive performance[10]. Empirical evidence from clinical prediction studies further demonstrates that ensemble often perform better than individual learners and exhibit greater stability under repeated resampling assessments[11, 12].

These approaches frequently limit variability by staying inside particular learning frameworks. This led to an interest in stacking, which leverages the benefits of several learners by combining predictions from different algorithms in a secondary learning process. The Super Learner method, which ideally employs cross-validation and ensemble weighting for enhanced prediction performance, is an example of stacking.

### 1.2. Development of the Super Learner Framework

By integrating many learning methods, the idea of stacking, also known to as stacked generalization was presented as a method for enhancing prediction performance. Stacking incorporates predictions from many algorithms into a single predictive model, in contrast to conventional ensemble approaches that combine comparable base learners. The fundamental justification is that many algorithms may capture different facets of the data-generating process, and their combination might produce predictions that are more reliable and accurate than those made by any one learner alone[13, 14].

Polley and van der Laan (2010) presented the Super Learner framework, which builds on layered generalization. Candidate learners are combined using convex weights chosen through risk reduction in this cross-validation-based ensemble approach. Their work demonstrated that the Super Learner always performs as well as or better than the best individual learner and provided empirical support for its oracle property[15].

The Super Learner stands out due to its solid theoretical framework. The Super Learner has the oracle property, which asserts that its predictive performance asymptotically converges to that of the optimal convex combination of candidate learners under very modest regularity conditions. Simply put, this means that the Super Learner will always beat any single constituent algorithm and will perform at least as well as the top learner in its library as sample size increases. The method’s increasing popularity across several application areas has been greatly aided by this theoretical guarantee[15].

Future developments extended the theoretical foundations of the Super Learner. In contrast to conventional global learner selection techniques, Valdes et al. (2022) presented the Conditional Super Learner, which carries out learner selection conditionally across parts of the covariate space and showed positive convergence qualities. These developments emphasize the significance of learner selection stability and oracle behavior in ensemble frameworks[16].

Prediction problems in healthcare have seen an increase in the use of the Super Learner framework. Applications include prediction of severe COVID-19 outcomes among cardiovascular patients[17], risk prediction of long COVID [18], cardiovascular event prediction following myocardial infarction[19], and diabetic kidney disease prediction among newly diagnosed patients with type 2 diabetes [20]. Super Learner continuously equaled or outperformed the performance of individual machine learning algorithms throughout these investigations.

### 1.3. The Role of Data Preprocessing in Machine Learning

It is commonly acknowledged that data preparation is an essential part of machine learning processes. Data cleaning, transformation, normalization, standardization, feature engineering, and feature selection are common preprocessing techniques that enhance data quality and predictive performance [21, 22].

Preprocessing decisions can have a significant impact on downstream analytical results, as several investigations have shown. While Amato and Di Lecce (2023) found that preprocessing procedures like normalization and dimensionality reduction considerably changed clustering results, Rahman (2019) found that z-score normalization performed better than alternative normalizing techniques as data size rose. In a similar vein, Wanyonyi and Masinde (2025) discovered that data transformation and feature selection significantly improved the performance of several machine learning algorithms [23–25].

However, while certain machine learning techniques are immune to scaling variations, not all of them call for preprocessing. Therefore, using these methods arbitrarily might make calculations more difficult without offering many advantages. Additionally, there is not enough clarity about the need for preprocessing in ensemble frameworks, which emphasizes how crucial it is to assess its effects in order to create effective machine learning procedures.

### 1.4. Preprocessing-Invariant Learners and Emerging Evidence

The algorithm determines how successful preprocessing is in machine learning. Tree-based algorithms, like CART, are frequently regarded as preprocessing-invariant as they rely on recursive partitioning rather than numerical scales, even though many methods benefit from feature scaling. As a result, tree-based models often require less preprocessing than techniques like support vector machines and neural networks. Preprocessing can improve predictive performance, especially for distance-based approaches, according to studies like those by Misra and Yadav (2019) [26] and Martinović et al. (2026) [27].

Preprocessing, which includes methods like normalization and feature selection, is frequently incorporated into the modeling pipeline in Super Learner or stacked ensemble research. None, however, has investigated the potential effects of preprocessing on learner weights, oracle behavior, or stability in an ensemble setting. Preprocessing may not affect individual tree-based models, but the interaction within a weighted ensemble may change depending on how well candidate learners perform during crossvalidation [19, 20, 28–30].

Preprocessing-invariant learners are common, but little is known about how they affect Super Learner ensembles, especially when it comes to learner weight distribution and oracle performance. This suggests that preprocessing in strong ensemble systems has to be evaluated more thoroughly.

### 1.5. Research Gap and Study Motivation

Little is known about how preprocessing affects Super Learner’s internal behavior when it is made up of preprocessing-invariant learners, despite the fact that it has shown great predictive performance and theoretical optimality. The majority of current research has concentrated on prediction accuracy, leaving significant issues with learner weight stability and oracle behavior unanswered. Therefore, the present study examines how preprocessing affects the theoretical characteristics and predictive performance of a Super Learner ensemble made up of learners that are invariant to preprocessing. Using three benchmark clinical classification datasets, namely the Pima Indian Diabetes, Heart Disease, and Indian Liver Patient datasets, the study evaluates the extent to which preprocessing influences predictive performance, learner weight allocation, and oracle behavior characteristics. By examining both predictive and theoretical aspects of ensemble learning, the study seeks to provide a more comprehensive understanding of the role of preprocessing in modern Super Learner frameworks. Specifically, the study seeks to:

1. Determine whether preprocessing significantly affects predictive performance of the Super Learner.
2. Determine whether preprocessing alters learner weight allocation within the Super Learner.
3. Assess whether preprocessing affects the oracle behavior of the Super Learner.

## 2. Materials and Methods

### 2.1. Study Design

The effect of preprocessing on the performance of a Super Learner ensemble composed of preprocessing-invariant learners was evaluated in this study using a comparative experimental design. Two conditions were examined using the framework: (i) a raw data condition without preprocessing and (ii) a preprocessed condition in which predictor variables were encoded and normalized prior to model training. The same Super Learner library and cross-validation approach were used in both training and validation processes for each dataset. Preprocessing was the sole variable.

For reliable estimations of ensemble behavior and predictive performance, the study used repeated cross-validation. To evaluate how preprocessing impacts prediction accuracy and the internal structure of the Super Learner framework, the investigation evaluated learner weight allocation and oracle behavior features in addition to conventional metrics.

### 2.2. Data Sources

Three publicly available benchmark healthcare datasets were used to evaluate the effect of preprocessing on Super Learner performance: the Pima Indians Diabetes Dataset [31], the Heart Disease Dataset [32], and the Indian Liver Patient Dataset (ILPD) [33]. These datasets were chosen because they are well-studied binary classification issues in medical and clinical prediction research, and they are commonly used to assess machine- and ensemble-learning methods.

The Pima Indians Diabetes Dataset, which is used for diabetes prediction, contains clinical and demographic information from female patients with Pima Indian heritage. While the Indian Liver Patient Dataset includes biochemical and demographic information from individuals with and without liver illness, the Heart illness Dataset includes measures pertaining to the presence or absence of cardiovascular disease. These datasets offer a variety of contexts for assessing preprocessing effects and strengthening the generalizability of findings since they differ in sample size, predictor makeup, and illness characteristics.

Before model development, each dataset was inspected for data quality issues, including missing values, duplicate observations, and inconsistencies in variable coding. Outcome variables were encoded as binary responses suitable for classification modeling.

Table 1 summarizes the key characteristics of the datasets used in this study.

**Table 1.** Characteristics of the datasets used in the study.

| Dataset | Observations ($n$) | Predictors ($p$) | Positive Cases (%) | Outcome |
| --- | --- | --- | --- | --- |
| Pima Indians Diabetes | 768 | 8 | 35 | Diabetes |
| Statlog - Heart Disease | 207 | 13 | 44 | Heart Disease |
| Indian Patient Liver Dataset | 583 | 10 | 71 | Liver Disease |

### 2.3. Experimental Workflow

The goal of the experimental workflow was to determine how preprocessing affected the Super Learner’s prediction performance. For a fair comparison, analyses were carried out on each dataset under two conditions: raw data and preprocessed data. All other elements of model construction were held constant.

The acquisition of data and quality evaluation were the first steps in the process, which was then divided into subsets for testing and training. While variables were normalized and encoded using training data in the preprocessed condition, original predictor values were utilized in the raw condition.

For both scenarios, the same base learner library was used to create a Super Learner ensemble. Predictive accuracy, learner weight allocation, and oracle behavior features were examined to determine the impact of preprocessing, and model performance was assessed using repeated cross-validation.

#### Experimental Workflow Framework

### 2.4. Data Partitioning and Resampling Strategy

To obtain reliable estimates of model performance, each dataset was partitioned into training and testing subsets using stratified random sampling to preserve the class distribution. The testing set was kept aside for final assessment, while the training set was utilized for cross-validation and model building. For a total of 100 resampling rounds, the model was trained using repeated 10-fold cross-validation ten times. To ensure that each fold functioned as a validation set throughout the process, the training data was divided into ten folds for each iteration, nine of which were utilized for training and one for validation. By lowering the variability of random sampling, repeated cross-validation yields more reliable estimates of prediction performance. This approach also enabled a thorough evaluation of learner weight stability across various validation scenarios.

To ensure that differences were only caused by preprocessing, identical resampling partitions were used for all comparisons between raw and preprocessed conditions.

### 2.5. Preprocessing Procedure

With our preprocessing-invariant learners, the contrast of the two scenarios constituted preprocessed, in which continuous predictors were normalized by centering around the mean and scaling by the standard deviation, and raw, in which predictor variables were unaltered.

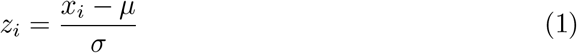

where (*x*_*i*_) denotes the original predictor value, (*µ*) represents the mean of the predictor calculated from the training data, and (*σ*) denotes the corresponding standard deviation.

Preprocessing parameters were only calculated from training data during each resampling iteration before being applied to testing and validation datasets in order to prevent information leakage. This method made guaranteed that model training was unaffected by validation or testing observations. The study assessed the benefits of preprocessing by comparing standardized and non-standardized data using the same learning algorithms and validation methods.

Figure 1 outlines the whole analytical workflow used in this investigation. The workflow provides a cohesive framework for addressing the study goals by demonstrating how each dataset advanced from preprocessing through model training and repeated cross-validation to the assessment of prediction performance, learner weight stability, and oracle behavior.

**Figure 1.**
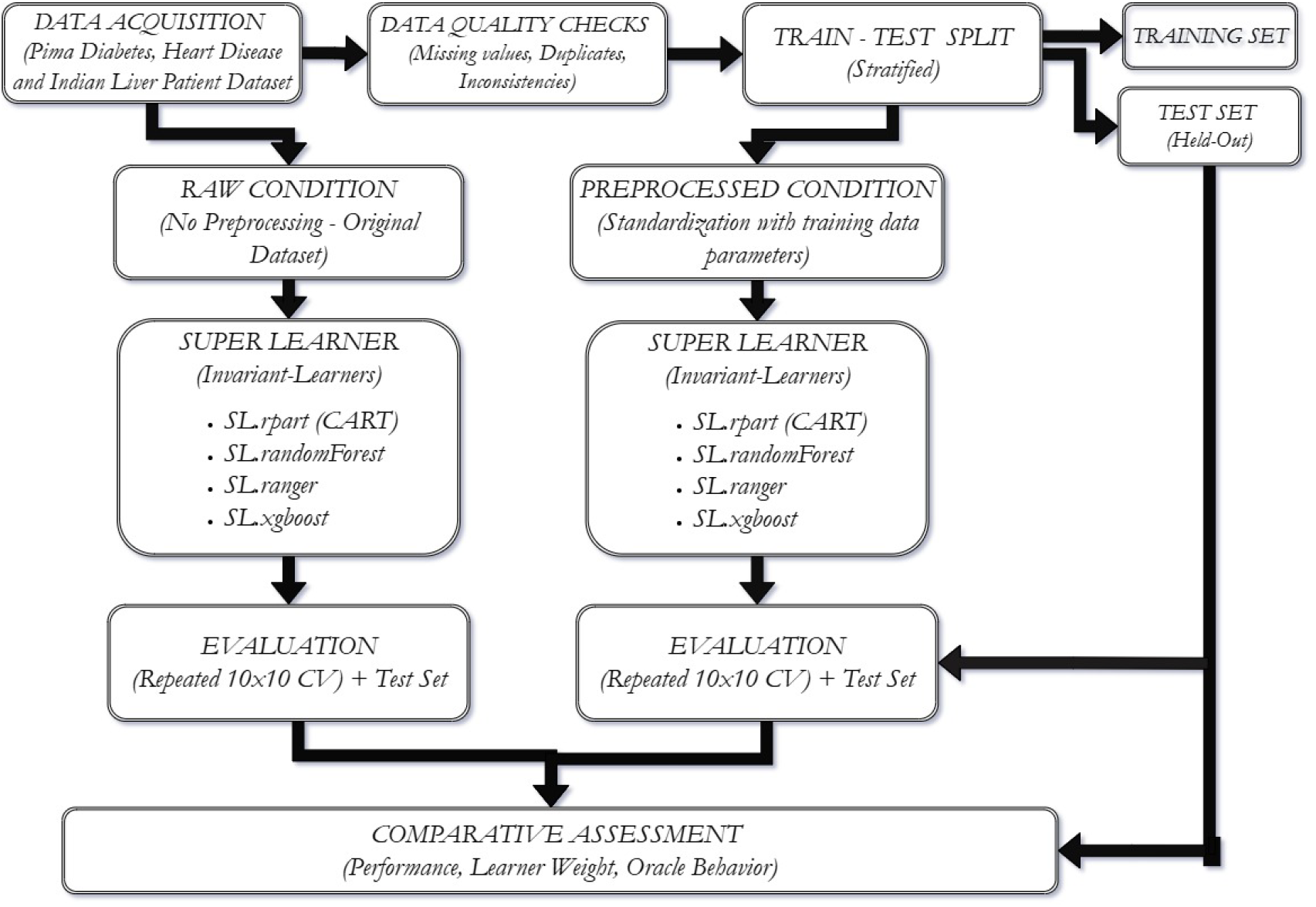
An outline of the experimental setup which encapsulates the whole analytical process.

### 2.6. Super Learner Specification

The main predictive modeling technique used in this study was the Super Learner framework. Super Learner is a stacking-based ensemble approach that uses cross-validated risk reduction to aggregate predictions from several candidate learners. The approach builds an optimally weighted ensemble to obtain predictive performance that is asymptotically equal to that of the best candidate learner or convex combination of learners, as opposed to choosing a single top-performing algorithm [15].

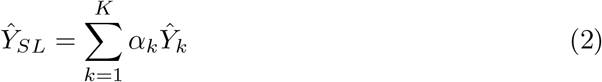

Constraints:

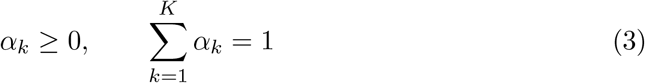

The Super Learner library consisted exclusively of preprocessing-invariant tree-based algorithms. Four candidate learners were included:

1. Classification and Regression Trees (CART; SL.rpart),
2. Random Forest (SL.randomForest),
3. Ranger Random Forest (SL.ranger), and
4. Extreme Gradient Boosting (SL.xgboost)

These learners were chosen because they are typically considered to be insensitive to feature scaling and normalization and are frequently employed in healthcare prediction research. As a result, they offer a suitable framework for assessing whether preprocessing affects ensemble behavior in spite of the constituent learners’ preprocessing invariance.

Repeated cross-validation was used to train candidate learners for each dataset and experimental condition. Non-negative least squares optimization (NNLS) was then used to aggregate the predictions produced by the candidate learners in order to estimate ensemble weights. The Super Learner prediction was the resultant weighted combination.

All datasets and preprocessing conditions used the same learner library, tuning parameters, and cross-validation approach.

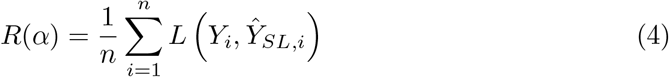

where *L*(*·*) denotes the loss function.

### 2.7. Predictive Performance Evaluation

Measures of discrimination, classification quality, and total prediction error were used to evaluate the impact of preprocessing on predictive performance. Each resampling iteration’s performance metrics were calculated and then compiled over all repetitions.

The Area Under the Receiver Operating Characteristic Curve (AUC), which gauges a model’s capacity to differentiate between positive and negative outcome classes over all potential classification thresholds, was used to assess discrimination ability [34].

The Matthews Correlation Coefficient (MCC) was used to evaluate the quality of the classification. In contrast to traditional accuracy metrics, MCC takes into account every component of the confusion matrix and offers a fair evaluation of predicted performance even when there is class imbalance [35].

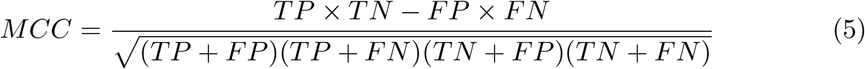

The Brier Score, which measures the difference between expected probability and actual results, was used to assess overall prediction inaccuracy. Better prediction accuracy and calibration are indicated by lower Brier scores [36].

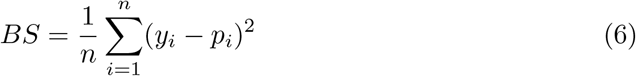

These metrics were chosen because, taken as a whole, they offer a thorough assessment of probability estimates, classification performance, and model discrimination.

### 2.8. Learner Weight Stability Analysis

Learner weight stability was assessed for each resampling iteration in order to determine whether preprocessing affects the Super Learner’s internal composition. The fitted Super Learner model was used to extract the ensemble weights allocated to the candidate learners for each fold.

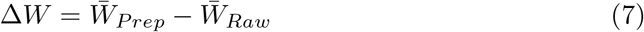

Weight stability was evaluated using a number of metrics. Initially, both raw and preprocessed mean learner weights were calculated. Second, to measure variations in learner contributions, average weight differences between the two scenarios were computed. Third, average ensemble weights were used to calculate learner ranks, and rank changes were analyzed to see if preprocessing changed the relative relevance of potential learners.

### 2.9. Oracle Behavior Analysis

Oracle-gap analysis and learner dominance measures were used to analyze oracle activity in order to determine if preprocessing affects the theoretical features of the Super Learner.

The candidate learner with the lowest cross-validated risk at a particular resampling iteration was designated as the oracle learner. The difference between the risk of the Super Learner and the risk of the top-performing prospective learner was used to compute the oracle gap:

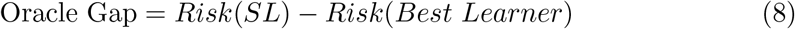

Stronger adherence to the Super Learner’s theoretical characteristics and a closer approximation of oracle performance are indicated by smaller oracle gaps [15, 37]

Oracle frequencies were determined for each learner by counting the number of resampling rounds in which a learner attained the lowest cross-validated risk, in addition to oracle-gap analysis. Learner dominance and whether preprocessing changed oracle selection patterns were evaluated using these frequencies.

To assess how much preprocessing impacts the Super Learner’s capacity to estimate oracle performance, the distribution of oracle gaps under raw and preprocessed circumstances was studied.

### 2.10. Statistical Analysis

R (Version 4.6.0) was used for all analyses. Descriptive statistics were used to describe learner weights, oracle behavior metrics, and predictive performance over resampling iterations. Because the resampling design was paired and the performance estimates were not normal, the paired Wilcoxon signed-rank test was used to compare raw and preprocessed conditions. The Benjamini–Hochberg false discovery rate approach was used to adjust p-values in order to account for multiple testing, and statistical significance was evaluated at the 5% level.

## 3. Results

### 3.1. Effect of Preprocessing on Predictive Performance

The predictive performance of the Super Learner for the Heart Disease, ILPD, and Pima datasets under both raw and preprocessed settings is shown in Table 2. The Matthews Correlation Coefficient (MCC), Brier Score, and Area Under the Receiver Operating Characteristic Curve (AUC) were used to evaluate the model’s performance.

**Table 2.** Comparison of predictive performance of the Super Learner under raw and preprocessed conditions across the three benchmark datasets.

| Dataset | Metric | Raw<br>(Mean ± SD) | Preprocessed<br>(Mean ± SD) | Mean Difference<br>(Preprocessed $-$ Raw) | Adjusted<br>$p$-value |
| --- | --- | --- | --- | --- | --- |
| Heart Disease | AUC | 0.901 ± 0.056 | 0.902 ± 0.055 | 0.0016 | 0.409 |
| | MCC | 0.648 ± 0.135 | 0.637 ± 0.137 | $-0.0110$ | 0.479 |
| | Brier Score | 0.127 ± 0.035 | 0.127 ± 0.037 | $-0.0001$ | 0.409 |
| ILPD | AUC | 0.746 ± 0.069 | 0.752 ± 0.065 | 0.0060 | **0.0017** |
| | MCC | 0.208 ± 0.133 | 0.219 ± 0.133 | 0.0105 | 0.282 |
| | Brier Score | 0.177 ± 0.023 | 0.175 ± 0.023 | $-0.0015$ | $< 0.001$ |
| Pima | AUC | 0.828 ± 0.041 | 0.829 ± 0.041 | 0.0013 | 0.256 |
| | MCC | 0.469 ± 0.093 | 0.472 ± 0.097 | 0.0025 | 0.495 |
| | Brier Score | 0.160 ± 0.019 | 0.160 ± 0.019 | $-0.0003$ | 0.409 |

The average performance under the preprocessed condition less the average performance under the raw condition was used to compute the mean difference. Whereas negative differences show better performance for the Brier Score, positive differences show better performance for AUC and MCC. The Benjamini–Hochberg false discovery rate method was used to produce adjusted $p$ -values. Results that are statistically significant are displayed in bold.

Preprocessing for the Heart Disease dataset resulted in minimal changes to any performance metrics. While the MCC slightly dropped from 0.648 ± 0.135 to 0.637 ± 0.137, the mean AUC slightly increased from 0.901 ± 0.056 to 0.902 ± 0.055. In the same manner, the Brier Score (0.127 ± 0.035 versus 0.127 ± 0.037) was essentially unaltered. After adjusting for multiple comparisons, none of these changes remained statistically significant.

Predictive performance for the ILPD dataset was somewhat improved by preprocessing. A statistically significant improvement was seen, with the mean AUC rising from 0.746 ± 0.069 to 0.752 ± 0.065 (adjusted p = 0.0017). Similarly, increased probabilistic prediction accuracy was shown by a drop in the mean Brier Score from 0.177 ± 0.023 to 0.175 ± 0.023. The change remained statistically significant even after multiple-testing correction (adjusted p *<* 0.001). Increase was not statistically significant, despite the MCC’s small rise from 0.208 ± 0.133 to 0.219 ± 0.133.

After preprocessing, only slight changes were found for the Pima dataset. The MCC rose from 0.469 ± 0.093 to 0.472 ± 0.097, and the mean AUC marginally increased from 0.828 ± 0.041 to 0.829 ± 0.041. In all scenarios, the Brier Score largely stayed the same. After accounting for multiple comparisons, none of these differences were statistically significant.

For the Heart Disease and Pima datasets, preprocessing generally had little effect on predicting accuracy. On the other hand, the ILPD dataset showed statistically significant gains in probability estimation and discrimination, indicating that the impact of preprocessing can vary depending on the dataset.

A visual evaluation of the variation in predicted performance over multiple cross-validation folds is given in Figure 2. Preprocessing yielded prediction performance that was generally stable throughout resampling iterations, as seen by the significant overlap between the raw and preprocessed distributions. The graphic shows that preprocessing had minimal impact on the overall dispersion and stability of the performance measures, with respect to the numerical summary shown in Table 2.

**Figure 2.**
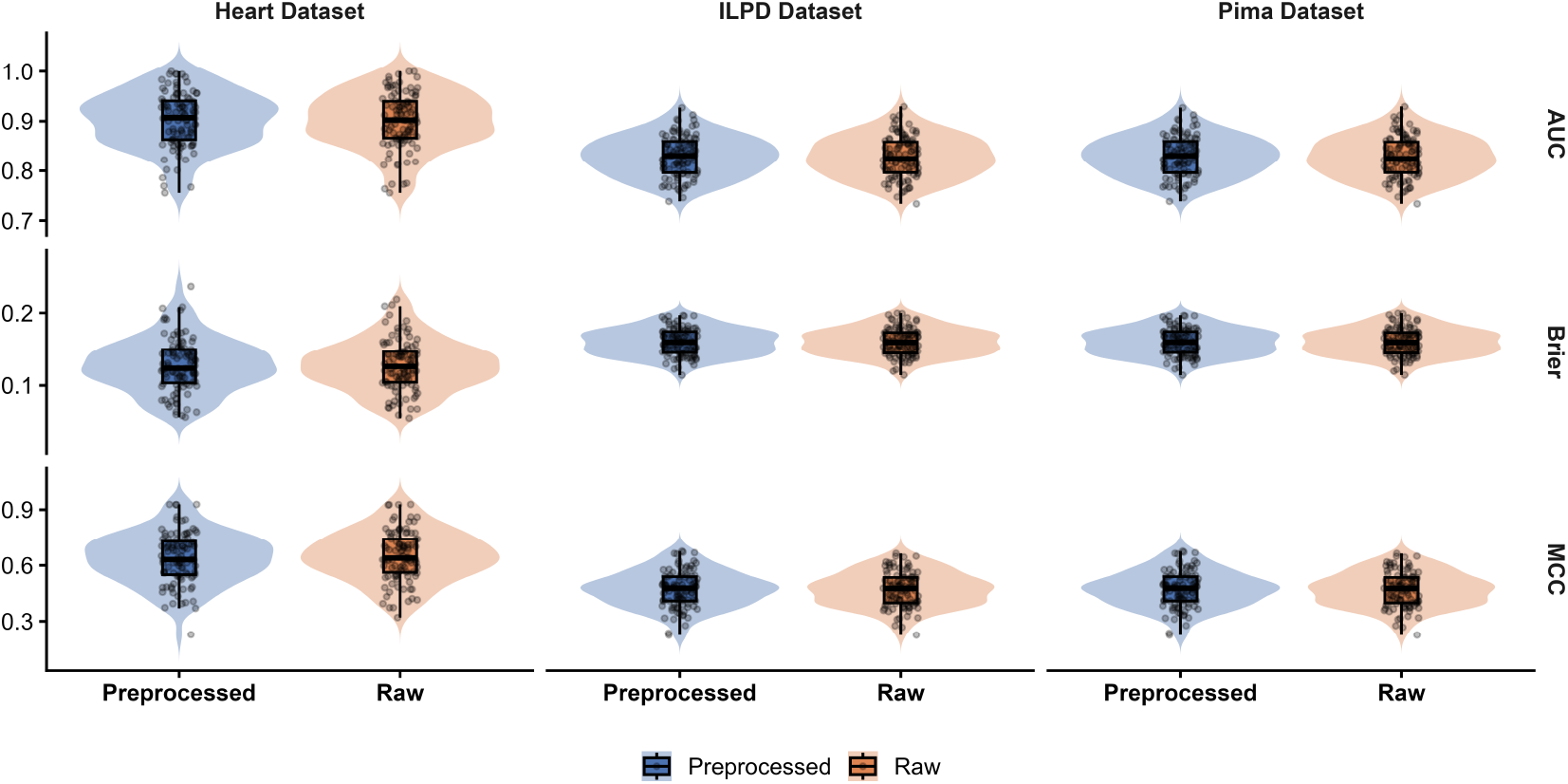
Super Learner prediction performance distribution in repeated cross-validation for the 3 datasets. With embedded boxplots showing the median and interquartile range, violin plots display the distribution of AUC, MCC, and Brier Score. Fold-specific performance estimations are indicated by individual points.

### 3.2. Effect of Preprocessing on Learner Weight Allocation

The average Super Learner weights assigned to each constituent learner under both raw and preprocessed settings across the three benchmark datasets are summarized in Table 3. Learner ranks, paired Wilcoxon signed-rank tests, and average ensemble weights were used to assess changes in learner significance.

**Table 3.** Comparison of Super Learner weight allocation under raw and preprocessed conditions across the three benchmark datasets.

| Dataset | Learner | Raw Weight | Prep Weight | $\Delta$ Weight | Rank | $\Delta$ Rank | Adj. $p$ |
| --- | --- | --- | --- | --- | --- | --- | --- |
| Heart Disease | RG | $0.438 \pm 0.287$ | $0.286 \pm 0.302$ | $-0.152$ | $1 \rightarrow 2$ | +1 | **0.005** |
| | XGB | $0.286 \pm 0.144$ | $0.274 \pm 0.153$ | $-0.012$ | $2 \rightarrow 3$ | +1 | 0.668 |
| | RF | $0.223 \pm 0.283$ | $0.362 \pm 0.316$ | $0.139$ | $3 \rightarrow 1$ | -2 | **0.006** |
| | CART | $0.052 \pm 0.079$ | $0.078 \pm 0.110$ | $0.025$ | $4 \rightarrow 4$ | 0 | 0.061 |
| ILPD | RF | $0.492 \pm 0.411$ | $0.552 \pm 0.418$ | $0.060$ | $1 \rightarrow 1$ | 0 | 0.569 |
| | RG | $0.480 \pm 0.411$ | $0.425 \pm 0.406$ | $-0.055$ | $2 \rightarrow 2$ | 0 | 0.569 |
| | CART | $0.015 \pm 0.036$ | $0.009 \pm 0.023$ | $-0.006$ | $3 \rightarrow 4$ | +1 | 0.418 |
| | XGB | $0.012 \pm 0.029$ | $0.014 \pm 0.035$ | $0.002$ | $4 \rightarrow 3$ | -1 | 0.668 |
| Pima | RF | $0.555 \pm 0.337$ | $0.501 \pm 0.360$ | $-0.053$ | $1 \rightarrow 1$ | 0 | 0.569 |
| | RG | $0.310 \pm 0.345$ | $0.367 \pm 0.361$ | $0.057$ | $2 \rightarrow 2$ | 0 | 0.569 |
| | CART | $0.135 \pm 0.097$ | $0.131 \pm 0.090$ | $-0.004$ | $3 \rightarrow 3$ | 0 | 0.695 |
| | XGB | $0.000 \pm 0.002$ | $0.000 \pm 0.004$ | $0.000$ | $4 \rightarrow 4$ | 0 | 0.787 |

Throughout the successive cross-validation folds, values are shown as Mean ± SD. The average change in learner weight after preprocessing (Preprocessed – Raw) is indicated by $\Delta$ Weight. Based on its average ensemble weight, the rank shows the learner’s place. RF = Random Forest; RG = Ranger; XGB = Extreme Gradient Boosting; CART = Classification and Regression Tree. The Benjamini–Hochberg false discovery rate method was used to produce adjusted $p$ -values.

Preprocessing significantly changed the learner weight distribution for the Heart Disease dataset. With its average weight rising from 0.223 ± 0.283 to 0.362 ± 0.316, Random Forest emerged as the dominating learner, improving ranking by two spots and producing a statistically significant difference after multiple-testing correction (adjusted p = 0.006). Ranger, on the other hand, saw the most weight loss, going from 0.438 ± 0.287 to 0.286 ± 0.302, along with a drop from first to second rank (adjusted p = 0.005). Both learner ranking and weight allocation showed relatively minimal improvements for XGBoost and CART.

After preprocessing, the learners’ relative relevance for the ILPD dataset was very consistent. In all scenarios, Random Forest maintained the greatest average ensemble weight, whereas Ranger continuously came in second. During the resampling procedure, CART and XGBoost were given very little weight. After accounting for multiple comparisons, none of the observed changes in learner weights were statistically significant.

Similarly, learner weight allocation for the Pima dataset was not significantly affected by preprocessing. CART and XGBoost made very little contribution to the ensemble, but Random Forest continued to be the leading learner, followed by Ranger. No statistically significant variations in learner weights were found, and learner ranks were maintained under both conditions.

For the ILPD and Pima datasets, preprocessing generally had little effect on learner weight allocation. The Heart Disease dataset, on the other hand, showed a redistribution of ensemble weights, mostly due to an increase in Random Forest’s contribution and a decrease in Ranger’s weight.

The distribution of ensemble weights among the constituent learners was affected by preprocessing, as shown in Figure 3. The image illustrates the direction and amount of these shifts, demonstrating that preprocessing mostly redistributed learner contributions while maintaining the overall ensemble structure, whereas Table 3 quantifies the average changes and statistical significance.

**Figure 3.**
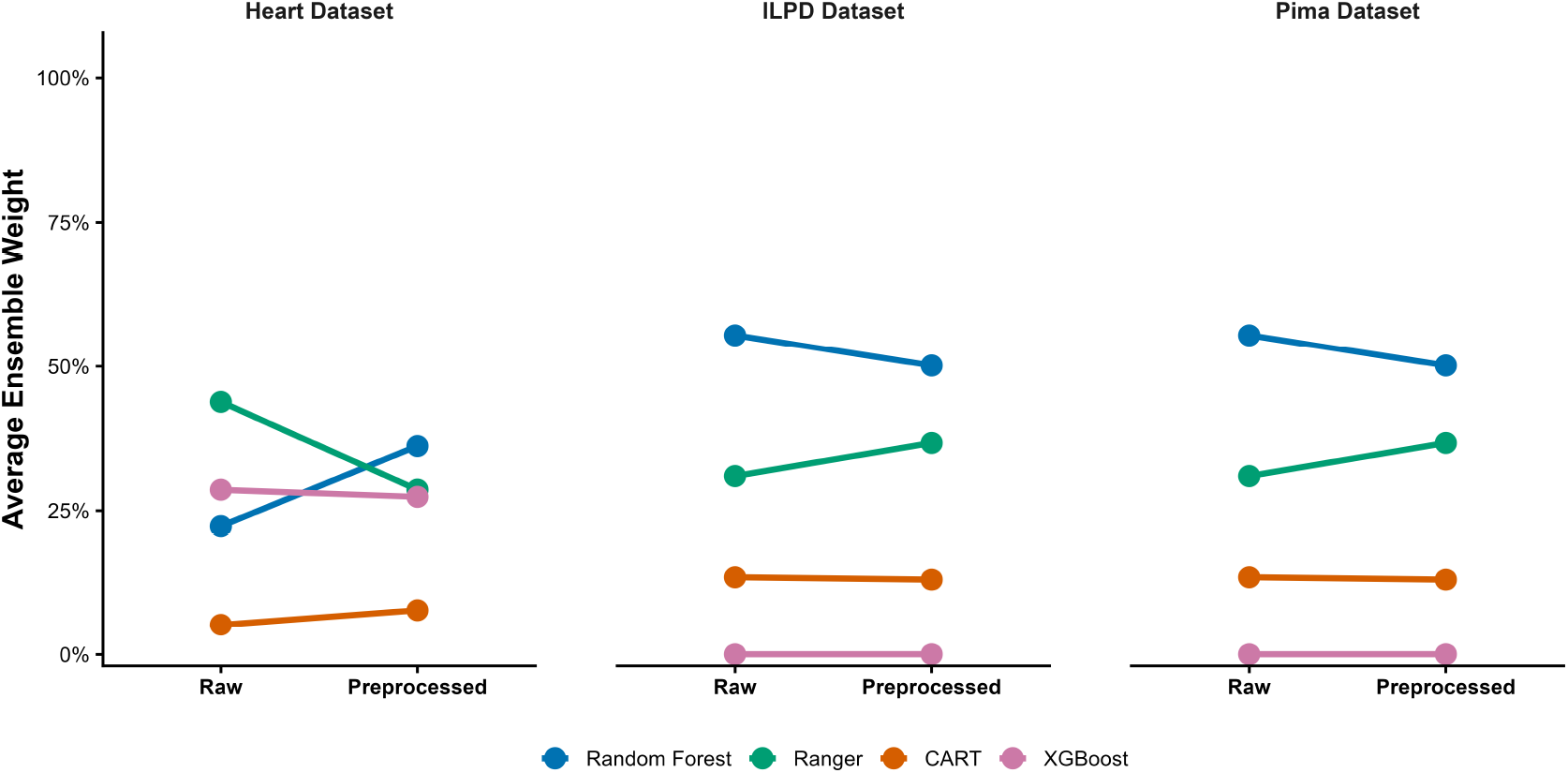
Average Super Learner weight distribution for the Heart Disease, Indian Liver Patient Dataset (ILPD), and Pima Indians Diabetes datasets under both raw and preprocessed settings. Each line connects the mean ensemble weight of a single base learner before and after preprocessing. The average learner contribution after preprocessing is shown by upward and decreasing curves, respectively.

### 3.3. Effect of Preprocessing on Oracle Behavior

The Oracle behavior of the Super Learner in both raw and preprocessed settings across the three benchmark datasets is summarized in Table 4. The Oracle Gap, which calculates the difference in predicting risk between the Super Learner and the top-performing candidate learner, was used to assess Oracle behavior.

**Table 4.** Comparison of oracle behavior of the Super Learner under raw and preprocessed conditions across the three benchmark datasets.

| Dataset | Raw Oracle Gap<br>(Mean ± SD) | Prep Oracle Gap<br>(Mean ± SD) | $\Delta$ Oracle Gap<br>(Prep $-$ Raw) | Dominant Oracle<br>(Raw → Prep) | Adj. $p$ |
| --- | --- | --- | --- | --- | --- |
| Heart Disease | 0.00080 ± 0.00233 | 0.00117 ± 0.00253 | 0.00037 | Ranger → RF | 0.615 |
| ILPD | 0.00097 ± 0.00071 | 0.00098 ± 0.00077 | 0.00001 | Ranger → RF | 0.979 |
| Pima | 0.00034 ± 0.00115 | 0.00011 ± 0.00112 | $-0.00024$ | RF → RF | 0.574 |

The gap in predicted risk between the Super Learner and the ensemble’s top-performing candidate learner is known as the Oracle Gap. The average change in Oracle Gap after preprocessing is represented by $\Delta$ Oracle Gap (Preprocessed $-$ Raw). The learner who is most commonly recognized as the oracle over several cross-validation folds is known as the Dominant Oracle. RF stands for Random Forest. The Benjamini–Hochberg false discovery rate method was used to produce adjusted $p$ -values.

The average Oracle Gap for the Heart Disease dataset rose somewhat after preprocessing, from 0.00080 ± 0.00233 in the raw condition to 0.00117 ± 0.00253. Nevertheless, after accounting for multiple comparisons, the observed difference was not statistically significant (adjusted p = 0.615). Additionally, preprocessing changed the dominant oracle learner; in the raw condition, Ranger was the most often chosen learner, but following preprocessing, Random Forest emerged as the dominating oracle.

Oracle Gap values for the ILPD dataset were almost the same in all scenarios, rising very slightly from 0.00097 ± 0.00071 to 0.00098 ± 0.00077. The associated difference (adjusted p = 0.979) was not statistically significant. After preprocessing, the dominant oracle learner changed from Ranger to Random Forest, but the Oracle Gap’s size practically stayed the same.

In similar fashion, the Pima dataset showed extremely little Oracle Gaps in both scenarios. After preprocessing, the average Oracle Gap dropped from 0.00034 ± 0.00115 to 0.00011 ± 0.00112, although the change was not statistically significant (adjusted p = 0.574). In all scenarios, Random Forest continued to be the most effective oracle learner.

Overall, preprocessing had minimal impact on the Super Learner’s oracle approximation for all three datasets. The Heart Disease and ILPD datasets showed changes in the dominant oracle learner, but these changes were not accompanied by statistically significant differences in Oracle Gap, suggesting that the Super Learner’s theoretical oracle behavior was largely maintained after preprocessing.

The fold-to-fold fluctuation in Oracle Gap under both raw and preprocessed circumstances is shown in Figure 4, which enhances the Oracle summary in Table 4. Preprocessing did not cause significant changes in oracle behavior, as seen by the near concentration of values around zero throughout numerous cross-validation folds, which shows constant oracle approximation.

**Figure 4.**
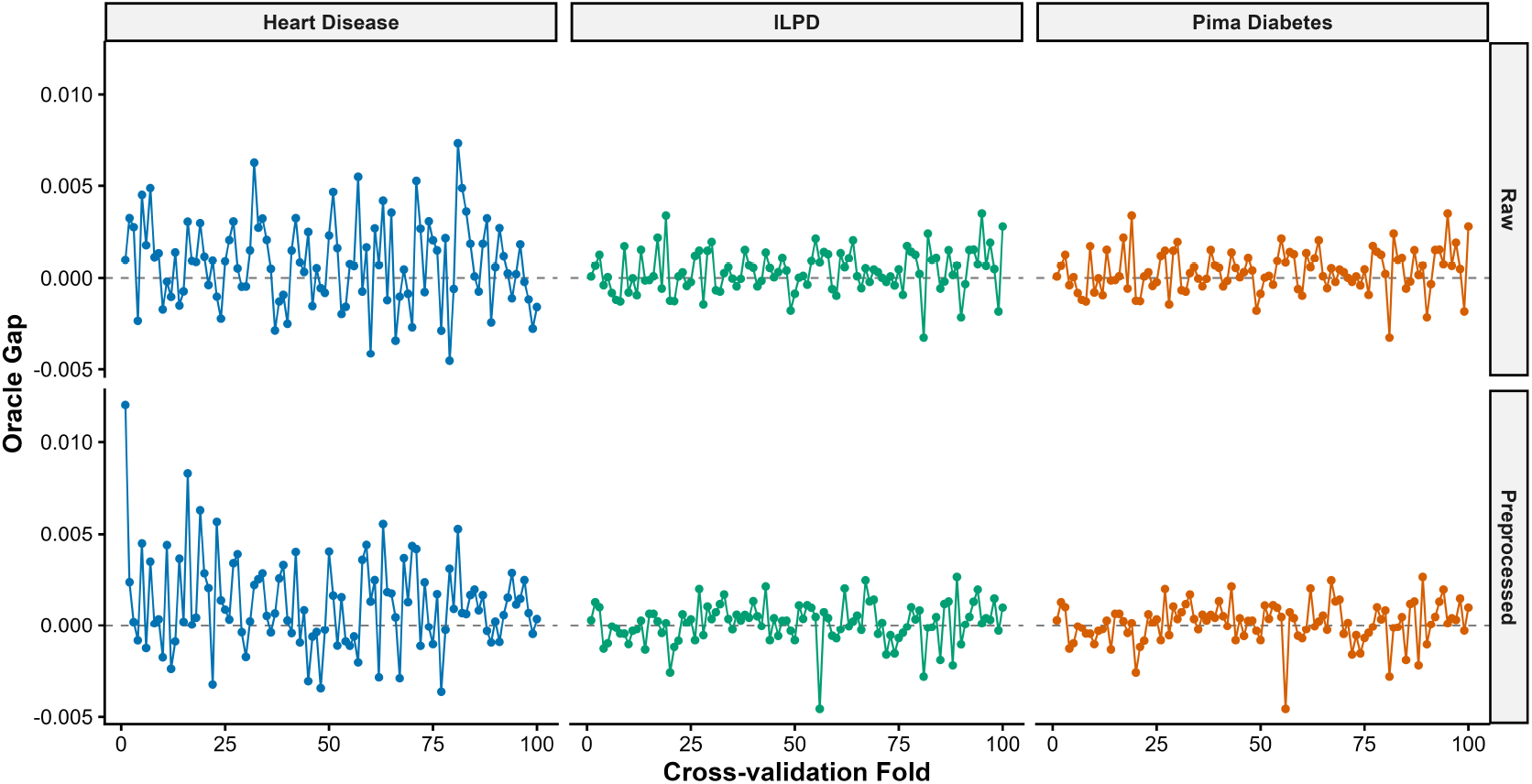
Oracle Gap for Pima Indians, Indian Liver Patient Dataset (ILPD), and Heart Disease dataset across cross-validation folds is displayed. Negative numbers imply a lesser risk, but positive values reflect that the Super Learner had a higher expected risk than the top learner. Oracle performance is closely approximated by values close to zero.

## 4. Discussion

### 4.1. Overview of the Study

This study examined whether preprocessing affects the Super Learner’s internal behavior and prediction performance when the ensemble library only includes preprocessing-invariant tree-based methods. Overall, across the three benchmark clinical datasets, preprocessing had no effect on oracle behavior, learner weight allocation, and predictive performance. The ILPD dataset showed statistically substantial improvements in AUC and Brier Score, but the Heart Disease and Pima datasets showed very slight changes. These results imply that rather than being a general requirement for tree-based Super Learner ensembles, the impact of preprocessing is mostly dictated by dataset features.

The theoretical properties of tree-based learning algorithms discussed by Misra and Yadav (2019) and Martinović et al. (2026), who showed that algorithms based on recursive partitioning are typically insensitive to feature scaling because their splitting criteria depend on the ordering of predictor values rather than their numerical magnitudes, are consistent with the limited improvements in predictive performance seen for the Heart Disease and Pima datasets. Therefore, it is doubtful that standardization and normalizing would significantly alter the prediction rules generated by CART, Random Forest, Ranger, or XGBoost. Preprocessing may still be helpful for datasets with varied predictor distributions or higher numerical variability, though, as seen by the statistically significant improvement seen for the ILPD dataset. The results of Rahman (2019), Amato and Di Lecce (2023), and Wanyonyi and Masinde (2025), who found that depending on the properties of the underlying data, suitable preprocessing techniques can enhance predictive performance, are supported by this observation.

Beyond predictive accuracy,this study looked at how preprocessing affects learner weight distribution, a topic that has gotten very little attention in the Super Learner literature. Without examining how preprocessing influences the internal distribution of learner weights, prior Super Learner applications have mostly focused on increases in predictive accuracy across biomedical applications, such as COVID-19 prognosis, cardiovascular risk prediction, and diabetic kidney disease prediction. The current findings show that learner weights for the ILPD and Pima datasets were relatively steady, while only the Heart Disease dataset had a redistribution of weights, with Random Forest taking the position of Ranger as the dominating learner. The overall predictive performance was barely altered in spite of this shift. This result emphasizes that learner weight allocation offers supplementary information beyond traditional performance measures and implies that preprocessing may change the ensemble’s internal composition without significantly altering its exterior prediction performance. The Oracle study also showed that preprocessing had little effect on the Super Learner’s theoretical behavior. Oracle Gaps were consistently tiny across all three datasets, and there were no statistically significant differences between the preprocessed and raw circumstances. The Super Learner asymptotically performs at least as well as the best candidate learner or the optimal convex combination of learners inside its library, according to Polley and van der Laan’s oracle property, which is empirically supported by these results. For the Heart Disease and ILPD datasets, Random Forest replaced Ranger as the predominant oracle learner, however the Oracle Gap’s size hardly altered. This implies that despite maintaining the Super Learner’s capacity to approach oracle performance, preprocessing may have an impact on which learner is chosen as the oracle. Additionally, by showing that oracle approximation is constant even when preprocessing changes the identity of the dominant learner, our results supplement the work of Valdes et al. (2022), who focused on learner selection and oracle behavior inside Super Learner frameworks.

This study’s extension of earlier preprocessing research from individual machine learning algorithms to stacked ensemble learning is a significant addition. While Super Learner studies have typically treated preprocessing as a standard part of the modeling pipeline without specifically looking at its impact on learner weights or oracle behavior, existing preprocessing studies have primarily assessed its impact on isolated classifiers or clustering algorithms. This study offers a more thorough understanding of how preprocessing impacts ensemble learning by concurrently assessing prediction performance, learner weight distribution, and oracle behavior. The findings show that preprocessing has no effect on the theoretical characteristics of the Super Learner and only slightly improves predictive accuracy when the candidate library is made up entirely of preprocessing-invariant learners.

Practically speaking, these results imply that when building Super Learner ensembles made up solely of preprocessing-invariant tree-based algorithms, normal preprocessing might not necessarily be required. Without jeopardizing prediction accuracy or oracle behavior, removing pointless preprocessing stages can streamline modeling procedures, save computing load, and increase reproducibility. However, the modifications noted for the ILPD dataset suggest that preprocessing choices should be informed by the dataset’s features rather than being applied arbitrarily. To ascertain whether the findings of this study apply to a wider range of machine learning scenarios, future research should examine heterogeneous Super Learner libraries that integrate preprocessing-sensitive and preprocessing-invariant algorithms, as well as high-dimensional biomedical datasets, survival outcomes, and multiclass classification problems.

## Data Availability

All data produced in the present work are contained in the manuscript and are publicly available online.

https://www.kaggle.com/datasets/kumargh/pimaindiansdiabetescsv

https://archive.ics.uci.edu/dataset/145/statlog+heart

https://archive.ics.uci.edu/dataset/225/ilpd+indian+liver+patient+dataset

## Acknowledgements

The authors would like to thank the developers and maintainers of the publicly available benchmark datasets used in this study. We also acknowledge the R development team and the contributors of the open-source packages that facilitated the implementation of the Super Learner framework and the statistical analyses conducted in this research.

## Disclosure statement

The authors declare that there are no competing interests or conflicts of interest regarding the publication of this manuscript.

## Data Availability

All datasets supporting this study has been included in the article.

## Funding

This research received no external funding.

## Authors Contributions

RD and DD were responsible for conceptualization and the study design. NKOA, YAB, KAS,ROA and JW contributed to the writing of the manuscript. RD and RAD performed the data visualization and analysis. RD and DD drafted the manuscript. All the authors read and approved the final manuscript.

## Notes

### Competing Interest Statement

The authors have declared no competing interest.

### Author Declarations

Three publicly available benchmark healthcare datasets were used in the study: 1. Pima Indians Diabetes: https://www.kaggle.com/datasets/kumargh/pimaindiansdiabetescsv 2 Heart Disease Dataset: https://archive.ics.uci.edu/dataset/145/statlog+heart 3. Indian Liver Patient Dataset: https://archive.ics.uci.edu/dataset/225/ilpd+indian+liver+patient+dataset

